# ParkProReakt - Evaluation of a proactive approach to health care in Parkinson’s disease: a study protocol for a randomised controlled trial

**DOI:** 10.1101/2024.11.29.24318185

**Authors:** Marlena van Munster, Johanne Stümpel, Anna J Pedrosa, Kati Niemand, Ingmar Wellach, Dirk Becker, Isabel Doblinger, Kristina Schmidt, René Reiners, Daniel Wolferts, Anika Martin, Marius Nisslmüller, Keywan Sohrabi, Volker Groß, Birgit Samans, Patrick Fischer, Mohammed Osman Ashraf, Rebecca Wichratz, Marcin Grzegorzek, Xinyu Huang, Artur Piet, Mona Irsfeld, Christian Trense, Paulina M. Olgemöller, Ümran Seven, Ann-Kristin Folkerts, Elke Kalbe, Hannes Böbinger, Jens Dapper, Lars Wohlfahrt, Max Geraedts, Natalie Altschuck, Linda Kerkemeyer, Carsten Eggers, David J Pedrosa

## Abstract

**Introduction:** Parkinson’s disease (PD) causes significant impairment due to both motor and non-motor symptoms, which severely impact patients’ health-related quality of life (HRQoL) and increase caregiver burden. Given the rising prevalence of PD in an aging population, particularly in Germany, the need for innovative and resource-efficient healthcare approaches is paramount. The complexity of PD symptoms and the necessity for individualised, multidisciplinary and digital health technology-based care are widely acknowledged; however, access to specialist care remains limited, particularly in rural areas. Current healthcare systems are frequently ill-equipped to deliver timely, personalised interventions. In response to these challenges, the ParkProReakt project aims to enhance PD care through a proactive, technology-enabled, multidisciplinary approach designed to improve patient HRQoL and alleviate caregiver burden.

**Methods and analysis:** A randomised controlled trial will assess the efficacy and cost-effectiveness of ParkProReakt - a proactive, multidisciplinary, digitally supported care model for community-dwelling people with Parkinson’s disease (PwPD) - compared with standard care. We will recruit a total of 292 PwPD and their informal caregivers living in two diverse regions in Germany. The primary outcome measure will be patients’ HRQoL as measured by the PDQ-39, obtained at baseline, monthly and at completion of participation. Secondary outcomes comprise patients’ subjective wellbeing, incidence or change of long-term care needs, global cognition and disease progression, utilisation of health care services including hospitalisations, caregiver burden and health care costs. Statistical analysis will include t-tests for HRQoL changes, GLM for confounders, and multilevel models for centre effects. Secondary outcomes and cost-effectiveness (ICER) will be analysed similarly, using R and SPSS.

**Ethics and dissemination:** The study protocol has been approved by the Ethics Committees of the Medical Associations of Hesse and Hamburg. The results of our study will be reported to the funding body and disseminated through scientific publications and presentations at national and international conferences.

*Registration details:* This study was registered with the German Registry for Clinical Studies (DRKS) in both German and English - number: DRKS00031092.

*What is already known on this topic:* Parkinson’s disease imposes severe motor and non-motor challenges on patients, impacting their quality of life and caregiver well-being. The complexity of symptoms necessitates individualized, multidisciplinary, and digital health-based approaches to care. Despite a recognized need for proactive, scalable interventions in Parkinson’s disease care, existing health systems have limited capacity for implementing these comprehensive, resource-efficient models effectively.

*What this study adds:* This study introduces a novel, proactive, technology-based, patient-centered model of care for people with Parkinson’s disease, integrating wearable technology and an app to improve patient health-related quality of life. It rigorously assesses this model’s effectiveness and cost-efficiency in Germany.

*How this study might affect research, practice or policy:* The study’s findings could inform policy on proactive digital care for aging populations, improve Parkinson’s disease care accessibility, and offer a framework for chronic disease management using patient-centered, cost-effective, and multidisciplinary approaches.

## Introduction

Parkinson’s disease (PD) causes significant disability in patients due to motor and non-motor symptoms (1). As the disease progresses, decreases in Health-related quality of life (HRQoL) place a significant burden on patients and (informal) caregivers (2). With an ageing German society (3) and a growing prevalence of PD in the elderly, the number of people with PD (PwPD) will increase (4). This ongoing fundamental societal change requires the development of new and innovative but also resource efficient health care approaches to meet the multitude needs of PwPD.

Tailored medical therapies for PwPD during their progressive neurodegeneration become even more difficult by distinct phenotypes (5). Patients often report complex and fluctuating symptoms that require specialised, person-centred care (6). Experts agree that an individualised, comprehensive and multidisciplinary approach to care is essential and advocate for centralised care coordination and non-pharmacological interventions such as patient education and self-management support (7). Recent literature supports this view and calls for the incorporation of these aspects into novel care approaches for PwPD (8, 9).

In Germany, to date the responsibility for coordinating the care of PwPD lies primarily with neurologists and, in some cases, general practitioners. Substantial waiting times for appointments are common for PwPD in Europe (10), hindering the proactive, timely detection of potential complications that could prevent costly hospital admission (11). Poorer access to specialist care in rural areas was particularly striking during the COVID-19 pandemic (12, 13), which some may see as reflecting a more general deficiency in the health care system. Overall, modern approaches to personalised care by different professional groups, such as specialist nurses, remain the exception, so that valuable resources are wasted (14). Since the pandemic, there has been a growing awareness that technological advances can facilitate specialist consultation, promote the involvement of other professionals in the care process, and enable high quality care. However, while there has been an increasing focus on PD-specific technology in research, it has rarely been translated into clinical practice (15). There is an acute need for action to adapt the structure of care for PwPD (16), which calls for new and innovative approaches that provide comprehensive but also cost-effective care, taking into account the increasing complexity of therapies.

In 2015, Germany introduced the Innovation Fund (IF) to promote integrated care and health services research within the statutory health insurance system. Successful projects may transition into standard care following positive evaluation. The IF-funded ParkProReakt project aims at sustainably improving the care of PwPD. This shall be achieved by a cross-sectoral, proactive, demand-driven and technology-enabled care model. The objective of this study is to evaluate the efficacy of ParkProReakt through a randomised controlled trial (RCT) in order to assess the change in HRQoL, symptom severity and burden on informal carers compared to a standard care group.

### Objectives and hypotheses

With this project we propose a multidisciplinary, digitally supported care model for PwPD and their informal caregivers which will be evaluated in a randomised controlled trial (RCT). The central question of the scientific evaluation is whether the planned proactive integrated care model is associated with improvements in several aspects of care for PwPD. Specifically, the following research hypotheses will be addressed:

#### Primary

- PwPD participating in the ParkProReakt care model show better health-related QoL than those receiving standard care.

#### Secondary

- The subjective well-being of PwPD who participate in the ParkProReakt care model is higher than that of patients in standard care.
- Informal caregivers of PwPD participating in the ParkProReakt care model report less burden than those of patients receiving standard care.
- The ParkProReakt model of care results in reduced health care utilisation, particularly hospitalisation, among PwPD and thus leads to a reduction in disease-related costs.

For a detailed list of the registered study protocol cf. www.drks.de DRKS00031092./

## Methods and analysis

### Intervention

Our intervention involves the provision of a comprehensive care model over a six-month period by a multidisciplinary team of community nurses, PD nurses, telenurses and study physicians for movement disorders – all specially trained in the management of PwPD (see Figure 1 for an outline). Collaboration between team members is enabled by a digital infrastructure. The infrastructure consists of a web platform (care provider platform) for documentation of relevant patient data and communication between care providers, a smartphone application (app) and wearable sensors to monitor patient symptoms in their home environment, and a server-based data integration centre for storing data. The information collected from the PwPDs by the app and the wearables enable in-depth monitoring and includes regular assessments of HRQoL, patient-reported symptoms and motor tests (e.g. finger tapping). The telenurse conducts continuous reviews and evaluations of the monitoring data remotely, translating them into treatment requirements via the use of a status traffic light system. The system is designed to change colour (green, yellow or red) in response to pre-defined critical events. If symptoms worsen, the telenurse contacts the PwPD and initiates appropriate measures in collaboration with the PwPD and the multidisciplinary care team. Using the available data and close communication with the patient, the team can proactively initiate measures (e.g., medication change/reduction, telephone calls, consultation with the treating physician etc.) and exchange information on a multidisciplinary basis. Under the individualised approach, each patient follows his or her own personalised care pathway within our network, subject to detailed specifications.

**Figure 1:**
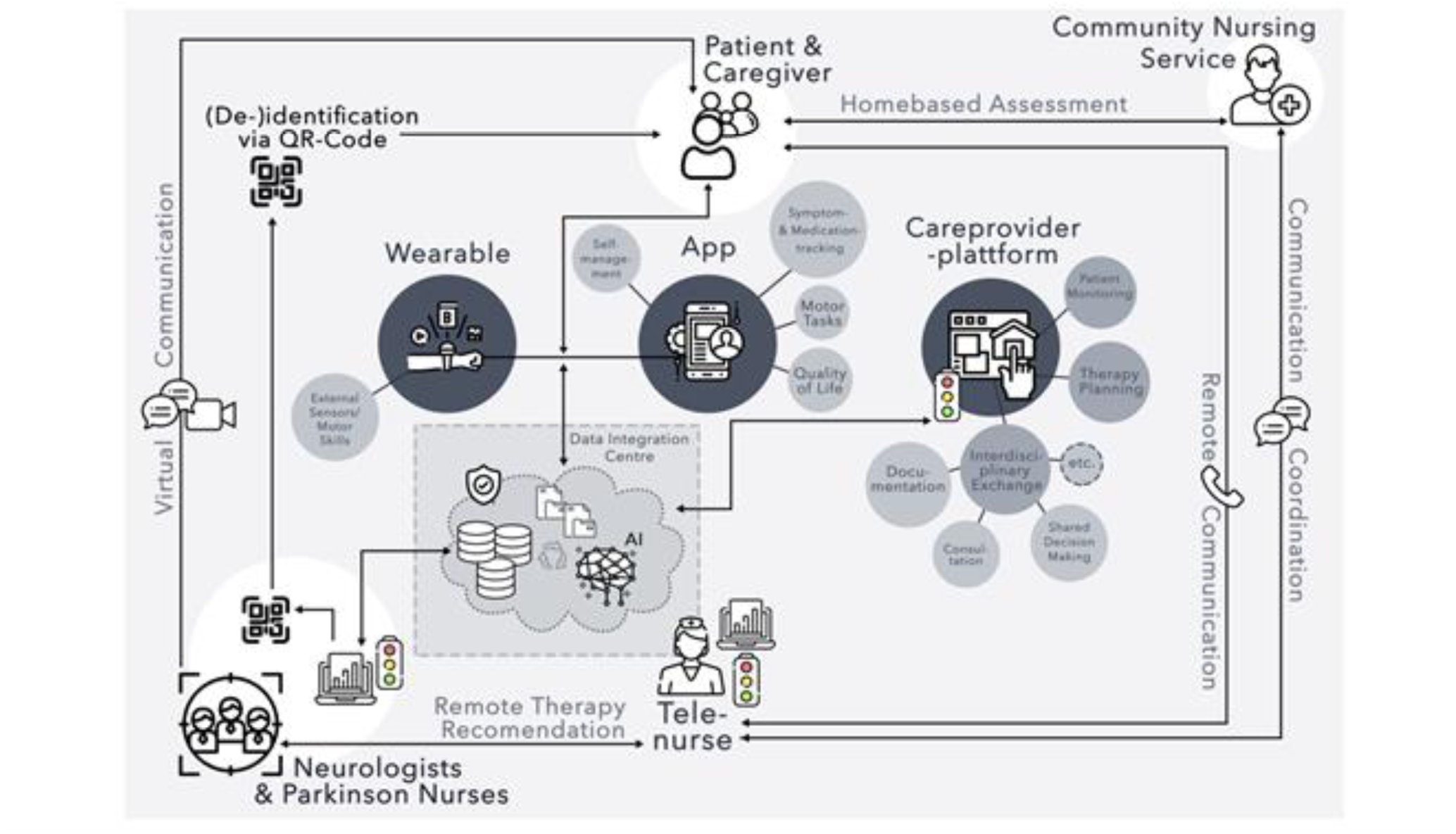
Outline of ParkProReakt (Icons: Flaticon.com)(17): At the heart of the complex intervention is the data integration centre, which is the key communication tool with patients and carers. Depending on the data collected via the app or wearables, different escalation levels of care are derived for the individual patient, which are delivered by a telenurse, a community nursing service, a Parkinson’s nurse and/or a neurologist. The care provider platform enables patient monitoring, multidisciplinary exchange and therapy planning.

The care provided includes three levels of care intensity, which PwPD enter, and exit based on identified care needs:

- Care level 1 involves continuous monitoring described above. In the event of clinical deterioration, relevant reduction of HrQoL or the occurrence of critical events that cannot be resolved by the telenurse, care will be intensified. Progressing to care level 2 or 3 depends on whether the need is considered by the telenurse to be primarily nursing or medical.
- PwPD with predominantly nursing needs are transferred to level 2 care, which involves a home visit by the community nursing service. The community nurses will take a thorough disease-specific nursing history, including a comprehensive nursing diagnosis. The documentation of the nursing history is reviewed by the multidisciplinary team. If there are any anomalies in the nursing history, the results are discussed with the study physician or at a regular nursing team meeting attended by at least one member of the three nursing groups. This may result in recommendations to patients, caregivers, physicians and care services not involved in the project. The relevant stakeholders, who are not directly involved in the study and do not have access to the project documentation, will be informed in writing. Patients and relatives receive advice by telephone from the telenurse, or in person from a community nurse if more appropriate and may be given educational material relevant to their situation.
- If the problems identified at care level 1 are judged to be strictly medical or have not been satisfactorily resolved by the measures at care level 2, the patient moves on to care level 3 (red traffic light). This highest level of care involves a phone or video consultation with the study physician. If care level 3 does not meet the patient’s needs, or if health problems are too severe to be managed in the community, hospital admission may be initiated. In the case of serious medical or nursing problems at the time of enrolment, direct transfer to level 2 or 3 may occur.In addition to the care provided by the multidisciplinary care team, PwPDs have the opportunity to find out about topics relevant to their illness and everyday life (exercise, nutrition or medication, cognition and psychosocial well-being) in an easy-to-understand way via a self-management tool in the smartphone app, which will be informed by qualitative interviews with both PwPDs and caregivers, designed by experts in the field, and pre-tested by PwPDs.

### Design

The whole study design is based on guidelines for complex health interventions (18) and includes the evaluation of change in clinical and economic outcomes via the here described RCT and an additional process evaluation for analysing the implementation of ParkProReakt (submitted). The outcome evaluation is based on a stratified (strata: study regions Hamburg / Marburg) randomised-controlled design, in which the degree of achievement of the primary and secondary outcomes of the intervention group is analysed in comparison to a standard care group. Given the obvious intervention and as it requires active cooperation, blinding takes place only at the level of the analyses but not for participants and care providers.

This protocol is reported according to the SPIRIT (Standard Protocol Items: Recommendations for Interventional Trials) guidelines (20), for details see also supplementary material 1.

### Setting

The study will be conducted in two structurally different German regions at two study centres: Praxis für Neurologie und Psychiatrie Hamburg Walddörfer (Hamburg, Northern Germany) and the Department of Neurology of the University Hospital of Marburg (Central Hesse, Central Germany). Hamburg, in northern Germany, is a large urban centre, whereas central Hesse, where Marburg is located, is more rural. These regional differences may influence general healthcare accessibility, patient demographics and care delivery. In this study, care will be provided in the home environment of PwPD, regardless of location.

### Participants

Participants must be adults (> 18 years) diagnosed with PD in stages 1 to 4 of the Hoehn & Yahr scale (as assessed by a physician), live in one of the designated study areas, have access to internet at home, have sufficient knowledge of the German language and be able to communicate. Family members/carers will be living or maintaining a close relationship with the patient and actively collaborating in his/her care. Nevertheless, PwPD may participate in the research project even if they do not have a caregiver or if the caregiver does not wish to participate. Patients cannot participate in the study if other cognition-impairing disease is present (e.g., severe dementia, tumors) or if comprehension and communication difficulties exist that interfere with inclusion criteria (e.g., ability to communicate verbally).

### Recruitment

Recruitment at both sites will follow a step-by-step procedure, re-evaluated every three months, and will be supported by the partner health insurance company (Techniker Krankenkasse, TK) as their involvement is integral to the Innofonds-sponsored nature of the study. Via the health insurance company’s database, TK’s insured can be reached and specifically addressed. Initially, all patients in the databases of the participating institutions who are insured with this major German health insurance company and who meet the inclusion criteria will be invited in writing for study participation. If the recruitment target is not met in the re-evaluation, the next escalation stage will be for the TK to invite its clients with a PD diagnosis who live close to the study sites to take part. In the final potential recruitment stage, volunteers listed in the Parkinson study database of the two study centres who are not insured with the TK will receive an invitation to the study. However, as there is evidence that only about 20% of patients can be motivated to participate in a trial, remaining in the first recruitment stage seems possible given the high number of PwPD in both locations (21).

### Sample Size

The calculation of the sample size was conducted at 80% power and a significance level of 5%. The PDQ-39 summed index (PDQ-39-SI) (22), was used as the primary outcome measure, which is a well-established tool for assessing the HRQoL (23). Previous literature has estimated clinically significant differences to be around five points (24, 25). It is worth mentioning that the PDQ-39-SI is widely employed for HRQoL assessments. The scale ranges from 0 to 100 and it is expected that PwPD fulfilling inclusion criteria have an average score of approximately 25±15 points on the HRQoL scale (26), resulting in an effect size for clinically significant differences of d = .33. This assumption is supported by research carried out by members of this group. They found that a simpler intervention, without digital assessments and self-management tools but consisting of an analogously coordinated integrated care model, resulted in substantial enhancements in HRQoL among a comparable population, with effect sizes of about d = .26 (21). Assuming the aforementioned conditions and an anticipated dropout rate of nearly 14% (21), the sample size calculation resulted in n = 146 participants in each group (intervention compared to control group) based on the independent samples t-test and heterogeneous variance.

### Randomisation

In both study regions, participants will be enrolled and then randomly assigned to either the intervention group (IG) or the control group (CG) using an allocation algorithm performed at study inclusion described previously (27). To account for known differences in certain disease characteristics and factors that may affect HRQoL, such as affective symptoms, disease severity, and gender, all enrolled individuals will be evaluated for these factors prior to randomisation. This evaluation includes an assessment of the individual’s gender, Hoehn & Yahr scale, and depression scores (measured with the BDI-II (Ref)). The algorithm employed is meticulously designed to achieve an optimal balance of potentially predictive factors across both groups, thereby minimising significant differences. By rigorously accounting for variables that may influence outcomes, this sophisticated allocation method ensures that participants in the intervention and control groups are as comparable as possible. It facilitates an equitable distribution of critical prognostic markers, significantly mitigating the risk of bias that could otherwise compromise the study results (27). Briefly, heterogeneity of the patient population has to be taken into account by an allocation according to certain rules, so that the two groups do not differ in these characteristics. For this purpose, a sequential calculation of a coefficient to reverse the most unequal distribution of prognostic markers will be used. It was shown in a simulation that equilibrium of predictive factors can be achieved (27).

### Enrolment

The review of the inclusion criteria and the random allocation to the groups will take place at a face-to-face appointment at the study centre by the respective team. Irrespective of randomisation, patients will be asked to complete a series of questionnaires as specified in below. Patients assigned to the IG will receive the digital technologies (wearable sensor and smartphone, Apple Inc., Cupertino, CA) required to deliver the care model and relevant instructions for usage. Training in the use of the technologies occurs in a face-to-face format; however, informational flyers will be distributed to the patients. Additionally, PwPD will be informed about access to the project’s website, where explanatory videos addressing any questions regarding the use of technology will be available (https://parkpro.parkinson-marburg.de/). The official start of the study is initiated by a follow-up phone call from the telenurse approximately one day after enrolment to ensure that all the technology is working and that the patient is aware of all the features. The CG receives standard health care only, which includes routine medical management by healthcare professionals, twice a year, pharmacological treatments in accordance with national guidelines (28) and access to supportive therapies such as physiotherapy, occupational therapy, and speech therapy, depending on individual needs. After 6 months, patients will be invited to the final study visit at their study centre, which will mark the end of their participation in the study. Dropouts and deviations will be monitored, and we will collect outcome data from those who discontinue whenever possible.

### Data collection

The primary outcome is the difference in change in HRQoL on the PDQ-39 sum index of PwPD participating in the ParkProReakt care model compared to those receiving standard care after the intervention period. Secondary outcomes, instruments used and frequency of assessment are shown in Table 1 whereas Table 2 provides an overview of the parameters specifically used for the cost-effectiveness analysis.

**Table 1:** Assessment of outcomes relevant to efficacy analysis during the 6-month study period.

| Parameter | Time of data collection |  |  | Assessment tool | IG* | CG* |
| --- | --- | --- | --- | --- | --- | --- |
| Primary outcome |  |  |  |  |  |  |
| HRQoL of patients | Baseline | monthly | 6-months | PDQ-39 (22, 29) | x | x |
| Secondary outcome |  |  |  |  |  |  |
| **Subjective wellbeing of the patients** | Baseline | monthly | 6- months | WHO-5 (30) | x | x |
|  | Baseline |  | 6- months | BDI-II (31) | x | x |
|  | Baseline |  | 6- months | EQ-5D-5L (32) | x | x |
| **Onset of long-term care needs or their change‡** | Baseline | 3- months | 6- months | Adapted version of the FIMA (33) | x | x |
| **Global cognition/ Disease progression** | Baseline |  | 6- months | MoCA (34) | x | x |
|  | Baseline |  | 6- months | MDS-UPDRS (35) | x | x |
|  | Baseline |  | 6- months | NMSS (36) | x | x |
| **Utilisation of health care services and hospitalisation frequency** | Baseline | 3- months | 6- months | Adapted version of the FIMA (33) | x | x |
|  | Continuously |  |  | provider documentation | x |  |
| **Caregiver burden** | Baseline |  | 6- months | ZBI (38) | x | x |
|  | Baseline |  | 6- months | BDI-II (31) | x | x |

IG\*: Intervention group; CG\*: Control group; ‡ The questionnaire was adapted to make it suitable for PwPD; PDQ: Parkinson's Disease Questionnaire; EQ-5D: EuroQol Health Questionnaire; WHO-5: Well-being Questionnaire; BDI-II: Beck's- Depression Inventory II; MoCA: Montreal Cognitive Assessment; MDS-UPDRS: Movement Disorder Society - Unified Parkinson's Disease Rating Scale; NMSS: Non-Motor Symptoms Scale; FIMA: Questionnaire for Health Services in Aging; ZBI: Zarit Burden Interview

**Table 2:** Assessment of outcomes relevant to cost-effectiveness analysis during the 6-month study period.

| Parameter | Time of data collection |  |  | Assessment tool | IG* | CG* |
| --- | --- | --- | --- | --- | --- | --- |
| **Utilisation of health care services and hospitalisation frequency** | Baseline | 3- months | 6- months | Adapted version of the FIMA (33) | x | x |
|  | Continuously |  |  | provider documentation | x |  |
| **(Total) costs** | Pre-observation period of 12 months | 3- months | 6 months | Statutory health insurance routine data of TK clients | x | x |

IG\*: Intervention group; CG\*: Control group; FIMA: Questionnaire for Health Services in Aging

All collected data are handled by the data integration centre of the University of Giessen during the intervention, pseudonymised and transferred to the data integration platform (DIP) in the form of FHIR resources. The collection of the various questionnaires to be filled out by the PwPDs and their caregivers is paper-based and will be digitised and pseudonymised transferred to the DIP using the “QForm” system. Double data entry of questionnaires is performed. All clinical scores and tests assessed through baseline or final study visits are handled the same way. Data access is restricted to care providers and the evaluation group and protected by passwords.

The data collection in ParkProReakt is divided into the project’s own care provider platform, the smartphone app and the wearable for capturing the motor tests. Data collected from the motor test via the wearable device (e.g., finger tapping) and the outcomes of the WHO-5 questionnaire, which assesses the HRQoL of the PwPD on a monthly basis through a smartphone application, alongside daily symptom reporting (also gathered via the application) enable patients’ direct reporting of non-emergency health events — such as relevant worsening of mobility and PD symptoms, sleep disturbances, vomiting, constipation, or pain — to the multidisciplinary health-care team. These data, in conjunction with real-time information from the wearable device and smartphone application, are presented on a platform accessible to healthcare providers enabling the multidisciplinary care team to review essential patient data (e.g., H&Y scale, medication regimes, etc.), document therapeutic decisions, and monitor nursing interventions. The dashboard records each activity, detailing timing and the individual responsible for its execution. These activities will later be made available to the evaluators in pseudonymised form. For the evaluation, the data is made available via the DIP. Access to the DIP is provided via a VPN connection to the Giessen research network. The evaluators’ personalised access points receive access to the pseudonymised data and can download them for further evaluation in the form of FHIR resources or CSV files.

### Data analysis

The primary endpoint will be analysed descriptively and inferentially with an independent samples t-test (group comparisons of changes in HRQoL in the IG and CG after the intervention period). Furthermore, General Linear Model (GLM) analyses will be conducted to investigate group differences at baseline which may affect the final outcome, as well as examine whether age, gender, comorbidities, or recruitment centre impacted the results. The flexible GLM framework permits additional modelling of multi-level data to assess both centres and their potential effects on the primary outcome. As the training of care providers involved in the intervention is centralised, the technical platform for both recruitment regions is supported by one project partner, and case discussions between health care professionals from both sites take place via telemedical consultation, no pronounced regional differences in care provision are expected. However, their potential effects are taken into account in the multivariate analysis models. Similar methods will be used to investigate analyses of secondary outcomes.

In the health economic evaluation, a cost-effectiveness analysis is planned to compare costs with effectiveness (incremental cost-effectiveness ratio, ICER). In this analysis, the cost per HRQoL gained will be determined. Therefore, the HRQoL (PDQ-39) will be used as the effect measure. The analysis of direct costs from a societal perspective considers the costs of the intervention and service use. For the main health economic analysis, the monetary valuation of health services (e.g., outpatient services, hospitalisation, medication, etc.) is performed using common valuation approaches (40, 41). In additional health economic analysis, routine data from the health insurance company TK are used to analyse the costs of insured persons who participated in ParkProReakt.

All quantitative data are analysed using statistical software such as R and SPSS.

### Patient and public involvement

The study design and care pathway were presented to and approved by a patient representative. In addition, patient representatives were actively involved in the development of the digital infrastructure, as well as in the formulation of the research question prior to the start of the project.

## Ethics and dissemination

Ethical approval for this study was obtained from the responsible state medical ethics committees in Hesse and Hamburg (Ref. 2022-3139-evBO and 2023-200762-BO-bet). Participants will sign a consent form stating that their decision to take part in the study is voluntary and that they can withdraw at any time. All personal information will be kept confidential as described above. The applicable data protection guidelines are followed and adhered to at all times. Participating PwPDs and their caregivers are informed about any use of their data and must consent to it.

Physical harm to PwPD is not anticipated as a result from participating in this study. However, participants may encounter potential burden associated with time constraints, both for themselves and their informal caregivers. The necessity to schedule study-related appointments and complete assessments may impose additional pressure on their regular routines, particularly for individuals with advanced PD. To address this concern, study visits will be arranged flexibly, and efforts will be made to minimise disruption to participants’ daily lives.

Regarding potential benefits, participants may experience an enhancement in their HRQoL through proactive, multidisciplinary, and digitally supported care. Informal caregivers may also benefit from the study’s approach, which aims to alleviate caregiver burden by providing support and monitoring the needs of PwPDs.

Results will be reported to the funding authority, presented at national and international conferences and disseminated through peer-reviewed publications. The findings will also be shared in close cooperation with the national Parkinson’s societies and patient organisations.

## Discussion

Demographic developments require administrations to devise effective neurological health care structures to address increasing demands (42). The World Health Organization has identified an increase in innovations of specialist support, such as telemedicine, to provide remote multidisciplinary assistance, as one of the significant steps to enhance PD care (43). Our proposed project aims to implement this concept for PwPD by transferring a complex but patient-centred care network into the German health care system. The efficacy, acceptability, and cost-effectiveness of this intervention will be assessed through a carefully designed RCT. The multidisciplinary care system is innovative and technically versatile. It was intentionally developed to be patient-friendly by incorporating advanced healthcare technologies like wearables and a smartphone app tailored to the needs of PwPD. Validated instruments are employed to assess HRQoL and symptom burden, ensuring the patient’s perspective is relevant to all care network processes. But the inclusion of affected individuals throughout the development process of our study was intended to maximise patient-centredness.

The use of patient-reported outcome measures (PROMs) offers several advantages. Firstly, intervention actions taken are relevant to patients. Secondly, patients benefit from being encouraged to participate in their own care. Thirdly, patient response rates are typically higher compared to those of clinicians. Fourthly, observer bias is avoided. Lastly, consideration of patient perspectives enhances public accountability of health services and professionals (44). In addition to using PROMs to capture patient perspectives objectively, we will also consider the relevance of the patient-carer dyad. The mental health implications for carers pose additional challenges to our health care system. Thus, our study protocol includes assessing the well-being of participating caregivers. Finally, the assessment is augmented with a cost-effectiveness examination of the utmost methodological quality, which is vital in an age of growing resource limitations in health care systems due to societal shifts.

In conclusion, we believe that our digitally supported multidisciplinary care network will effectively address the needs of patients and carers. However, our thorough evaluation study will also facilitate evidence-based decision making beneficial for clinicians and policy makers. The numerous evaluation findings are especially noteworthy in PD, where the perspectives of patients, carers and society warrants investigation.

## Trial Status

Protocol Version 1.0

The trial is currently in progress, with participant recruitment actively ongoing. Recruitment commenced in January 2024 and is anticipated to conclude by January 2025.

## Authors’ contributions

D.P. is the principal investigator, responsible for the overall quality and development of the protocol and ethical approvals. D.P. and M.v.M. are in charge of overseeing the project. C.E., D.P., M.v.M., I.W., D.B. were involved in the initial conception of the study. All authors contributed to refinement of the study protocol. M.G., N.A., L.K. were responsible for conceptualising the evaluation and will supervise the data collection as well as the analysis. D.P., J.S., M.v.M., A.P., I.W., D.B. will develop the implementation of the interventions. M.v.M., J.S. and A.P. wrote the first draft of the manuscript and all authors revised it critically for important intellectual content. All authors have read and approved the final manuscript.

## Funding statement

This work was supported by the Innovationfund of the German Federal Joint Committee (Innovationsfonds des Gemeinsamen Bundesausschuss G-BA); grant number [01NVF20019] and the Parkinson’s Foundation.

For information about the Innovation Fund:

Innovationsausschuss beim Gemeinsamen Bundesausschuss

P.O. Box 12 06 06

10596 Berlin

## Supporting information

Supplement 1 - Spirit Guideline

## Data Availability

All data produced in the present study are available upon reasonable request to the authors.

## References

1. Postuma RB, Berg D, Adler CH, Bloem BR, Chan P, Deuschl G, et al. The new definition and diagnostic criteria of Parkinson’s disease. The Lancet Neurology. 2016;15(6):546–8.

2. Klietz M, Schnur T, Drexel S, Lange F, Tulke A, Rippena L, et al. Association of motor and cognitive symptoms with health-related quality of life and caregiver burden in a German cohort of advanced Parkinson’s disease patients. Parkinson’s Disease. 2020;2020.

3. German Federal Statistical Office (Destatis). Bevölkerung in Deutschland 2023 [Available from: https://service.destatis.de/bevoelkerungspyramide/index.html#!y=2003&v=2.

4. Dorsey E, Sherer T, Okun MS, Bloem BR. The emerging evidence of the Parkinson pandemic. Journal of Parkinson’s disease. 2018;8(s1):S3–S8.

5. Eggers C, Pedrosa DJ, Kahraman D, Maier F, Lewis CJ, Fink GR, et al. Parkinson subtypes progress differently in clinical course and imaging pattern. 2012.

6. Storch A, Schneider CB, Wolz M, Stürwald Y, Nebe A, Odin P, et al. Nonmotor fluctuations in Parkinson disease: severity and correlation with motor complications. Neurology. 2013;80(9):800–9.

7. Bloem BR, Henderson EJ, Dorsey ER, Okun MS, Okubadejo N, Chan P, et al. Integrated and patient-centred management of Parkinson’s disease: a network model for reshaping chronic neurological care. The Lancet Neurology. 2020;19(7):623–34.

8. Lidstone SC, Bayley M, Lang AE. The evidence for multidisciplinary care in Parkinson’s disease. Expert Review of Neurotherapeutics. 2020;20(6):539–49.

9. Rajan R, Brennan L, Bloem BR, Dahodwala N, Gardner J, Goldman JG, et al. Integrated care in Parkinson’s disease: a systematic review and meta-analysis. Movement Disorders. 2020;35(9):1509–31.

10. Schrag A, Khan K, Hotham S, Merritt R, Rascol O, Graham L. Experience of care for Parkinson’s disease in European countries: a survey by the European Parkinson’s Disease Association. European Journal of Neurology. 2018;25(12):1410–e120.

11. Bloem, B R, Henderson, E J, Dorsey, E R, Okun, M S, Okubadejo, N, Chan, P, Andrejack, J, Darweesh S K L, Munneke, M. Integrated and patient-centred management of Parkinson’s disease: a network model for reshaping chronic neurological care. The Lancet Neurology. 2020; 19(7), 623–634.

12. van Munster M, Printz MR, Crighton E, Mestre TA, Pedrosa DJ, Consortium i-P. Impact of the COVID-19 pandemic on perceived access and quality of care in German people with parkinsonism. Frontiers in Public Health. 2023;11:1091737.

13. Wolff AW, Haller B, Demleitner AF, Pürner D, Niederschweiberer J, Cordts I, et al. Long-Lasting Impact of the COVID-19 Pandemic on Patients with Parkinson’s Disease and Their Relatives. Movement Disorders Clinical Practice. 2023;10(5):819–23.

14. van Munster M, Stümpel J, Thieken F, Ratajczak F, Rascol O, Fabbri M, et al. The Role of Parkinson Nurses for Personalizing Care in Parkinson’s Disease: A Systematic Review and Meta-Analysis. Journal of Parkinson’s Disease. 2022(Preprint):1–25.

15. Kleinholdermann U, Melsbach J, Pedrosa D. Remote-Messung bei idiopathischem Parkinson-Syndrom: Entwicklungen in Diagnose, Monitoring und Therapie. Der Nervenarzt. 2019;90(12).

16. Prell T, Siebecker F, Lorrain M, Eggers C, Lorenzl S, Klucken J, et al. Recommendations for standards of network care for patients with Parkinson’s disease in Germany. Journal of Clinical Medicine. 2020;9(5):1455.

17. Flaticon. 2023 [Available from: www.flaticon.com.

18. Möhler R, Köpke S, Meyer G. Criteria for reporting the development and evaluation of complex interventions in healthcare: revised guideline (CReDECI 2). Trials. 2015;16(1):1–9.

19. Ward J, Schaal M, Sullivan J, Bowen ME, Erdmann JB, Hojat M. The Jefferson scale of attitudes toward physician–nurse collaboration: A study with undergraduate nursing students. Journal of interprofessional care. 2008;22(4):375–86.

20. Chan A-W, Tetzlaff JM, Altman DG, Laupacis A, Gøtzsche PC, Krleža-Jerić K, et al. SPIRIT 2013 statement: defining standard protocol items for clinical trials. Annals of internal medicine. 2013;158(3):200–7.

21. Eggers C, Dano R, Schill J, Fink G, Hellmich M, Timmermann L. Patient-centered integrated healthcare improves quality of life in Parkinson’s disease patients: a randomized controlled trial. Journal of neurology. 2018;265:764–73.

22. Jenkinson C, Fitzpatrick R, Peto V, Greenhall R, Hyman N. The Parkinson’s Disease Questionnaire (PDQ-39): development and validation of a Parkinson’s disease summary index score. Age and ageing. 1997;26(5):353–7.

23. Stührenberg M, Berghäuser CS, van Munster M, Pedrosa Carrasco AJ, Pedrosa DJ, Consortium i-P. Measuring quality of life in Parkinson’s disease—A call to rethink conceptualizations and assessments. Journal of Personalized Medicine. 2022;12(5):804.

24. Peto V, Jenkinson C, Fitzpatrick R. Determining minimally important differences for the PDQ-39 Parkinson’s disease questionnaire. Age and ageing. 2001;30(4):299–302.

25. Horváth K, Aschermann Z, Kovács M, Makkos A, Harmat M, Janszky J, et al. Changes in quality of life in Parkinson’s disease: how large must they be to be relevant? Neuroepidemiology. 2017;48(1-2):1–8.

26. Butterfield LC, Cimino CR, Salazar R, Sanchez-Ramos J, Bowers D, Okun MS. The Parkinson’s Active Living (PAL) Program: a behavioral intervention targeting apathy in Parkinson’s disease. Journal of geriatric psychiatry and neurology. 2017;30(1):11–25.

27. Atkinson AC, Duarte BP, Pedrosa DJ, van Munster M. Randomizing a clinical trial in neuro-degenerative disease. Contemporary Clinical Trials Communications. 2023;33:101140.

28. Höglinger G., Trenkwalder C., et al., Parkinson-Krankheit, S2k-Leitlinie, 2023, in: Deutsche Gesellschaft für Neurologie (Hrsg.), Leitlinie für Diagnostik und Therapie in der Neurologie. Online: www.dgn.org/leitlinien (abgerufen am 01.10.2024)

29. Berger K, Broll S, Winkelmann J, Heberlein I, Müller T, Ries V. Untersuchung zur Reliabilität der deutschen Version des PDQ-39: ein krankheitsspezifischer fragebogen zur erfassung der lebensqualität von parkinson-patienten. Aktuelle Neurologie. 1999;26(04):180–4.

30. Brähler E, Mühlan H, Albani C, Schmidt S. Teststatistische prüfung und normierung der deutschen versionen des EUROHIS-QOL lebensqualität-Index und des WHO-5 wohlbefindens-index. Diagnostica. 2007;53(2):83–96.

31. Kühner C, Bürger C, Keller F, Hautzinger M. Reliabilität und validität des revidierten beck-depressionsinventars (BDI-II). Der Nervenarzt. 2007;78(6):651–6.

32. Hinz A, Kohlmann T, Stöbel-Richter Y, Zenger M, Brähler E. The quality of life questionnaire EQ-5D-5L: psychometric properties and normative values for the general German population. Quality of Life Research. 2014;23:443–7.

33. Seidl H, Bowles D, Bock J-O, Brettschneider C, Greiner W, Koenig H-H, et al. FIMA–Fragebogen zur Erhebung von Gesundheitsleistungen im Alter: Entwicklung und Pilotstudie. Das Gesundheitswesen. 2014:46–52.

34. Dalrymple-Alford J, MacAskill M, Nakas C, Livingston L, Graham C, Crucian G, et al. The MoCA: well-suited screen for cognitive impairment in Parkinson disease. Neurology. 2010;75(19):1717–25.

35. Goetz CG, Tilley BC, Shaftman SR, Stebbins GT, Fahn S, Martinez-Martin P, et al. Movement Disorder Society-sponsored revision of the Unified Parkinson’s Disease Rating Scale (MDS-UPDRS): scale presentation and clinimetric testing results. Movement disorders: official journal of the Movement Disorder Society. 2008;23(15):2129–70.

36. Jost W, Fuchs G, Reifschneider G, Odin P, Storch A, Ebersbach G. Validation of a German version of the NMSS (Non-Motor Symptom assessment Scale). Klinische Neurophysiologie. 2010;41(01):ID5.

37. Yahalom G, Simon E, Thorne R, Peretz C, Giladi N. Hand rhythmic tapping and timing in Parkinson’s disease. Parkinsonism & related disorders. 2004;10(3):143–8.

38. Bédard M, Molloy DW, Squire L, Dubois S, Lever JA, O’Donnell M. The Zarit Burden Interview: a new short version and screening version. The gerontologist. 2001;41(5):652–7.

39. Bowen GA. Document analysis as a qualitative research method. Qualitative research journal. 2009;9(2):27–40.

40. Bock J-O, Brettschneider C, Seidl H, Bowles D, Holle R, Greiner W, et al. Ermittlung standardisierter Bewertungssätze aus gesellschaftlicher Perspektive für die gesundheitsökonomische Evaluation. Das Gesundheitswesen. 2014:53–61.

41. Schöffski O. Grundformen gesundheitsökonomischer Evaluationen: Springer; 2007.

42. Feigin VL, Nichols E, Alam T, Bannick MS, Beghi E, Blake N, et al. Global, regional, and national burden of neurological disorders, 1990–2016: a systematic analysis for the Global Burden of Disease Study 2016. The Lancet Neurology. 2019;18(5):459–80.

43. World Health Organization. Parkinson Disease: A public health approach. Technical brief. Geneva, Switzerland; 2022.

44. Black N. Patient reported outcome measures could help transform healthcare. Bmj. 2013;346.

